# PSA/testosterone ratio as a potential biomarker to identify early progressors of adaptive therapy in metastatic castration sensitive prostate cancer

**DOI:** 10.1101/2025.05.08.25327179

**Authors:** Jill A. Gallaher, Robert A. Gatenby, Joel S. Brown, Alexander R. A. Anderson, Jingsong Zhang

## Abstract

**PURPOSE:** We aim to identify biomarkers of progression in an ongoing pilot trial using adaptive dosing in metastatic castrate sensitive prostate cancer (mCSPC).

**PATIENTS AND METHODS:** Men with mCSPC were given combined androgen deprivation therapy with an androgen receptor signaling inhibitor followed by a treatment break after achieving >75% PSA decline. This was followed by evolution-informed drug cycling to prevent resistance outgrowth. Just 6 of 16 patients have progressed at month 55, and we wish to identify predictive features that would help identify these patients. We compare clinical features, testosterone metrics, and PSA metrics over the trial cohort, and we fit a mathematical model to the dynamic patient-specific data to gain more mechanistic insight as to what might be driving early progression.

**RESULTS:** Significant clinical risk factors predicting progression included the risk status, the number of bone metastases, and the total number of metastases. However, analyses of the dynamical changes in PSA and testosterone during the initial cycle of treatment identified several useful features to discriminate early, late, and non-progressors. Specifically, a higher PSA at the end of the induction phase, a lower percent change in PSA over the induction period, and a higher average PSA/Testosterone ratio over the first cycle significantly predicted progressors. Thresholds for these metrics are able to distinguish the earliest progressors. A simple mathematical model coupling the PSA and the testosterone dynamics suggests a higher fraction of resistant cells exist prior to therapy for those that progress earlier.

**CONCLUSION:** PSA/Testosterone ratio is a useful feature to couple drug response to tumor burden dynamics and predict early progression from the first cycle of adaptive therapy.

## Introduction

We previously reported the primary endpoint of feasibility on an ongoing adaptive therapy study for metastatic castration sensitive prostate cancer (mCSPC) [NCT03511196] [1] . This trial follows a previous adaptive therapy trial for patients with metastatic castrate resistant disease (mCRPC), in which the cohort had received luteinizing hormone-releasing hormone (LHRH) treatment continuously and abiraterone adaptively to prolong treatment sensitivity and delay progression. In this mCSPC trial both LHRH analog and second-generation androgen receptor signaling inhibitors (ARSIs) like abiraterone are stopped after achieving more than 75% PSA decline from a total of 8-12 weeks of combined androgen deprivation therapy. At the time of off-treatment PSA or imaging progression, treatment is restarted and the choice of LHRH analog versus ARSI or both is based on the serum testosterone level at the time of progression. Treatment is stopped again after 50% or more PSA decline is achieved.

Now with a median follow up of 55 months, the secondary endpoints on median time to PSA progression and the median time to radiographic progression have not been reached. Six of the 16 enrolled patients have developed on treatment PSA progression (Figure 1). Here we compared the clinical features between progressors and non-progressors to identify biomarkers that will predict progression.

**Figure 1.**
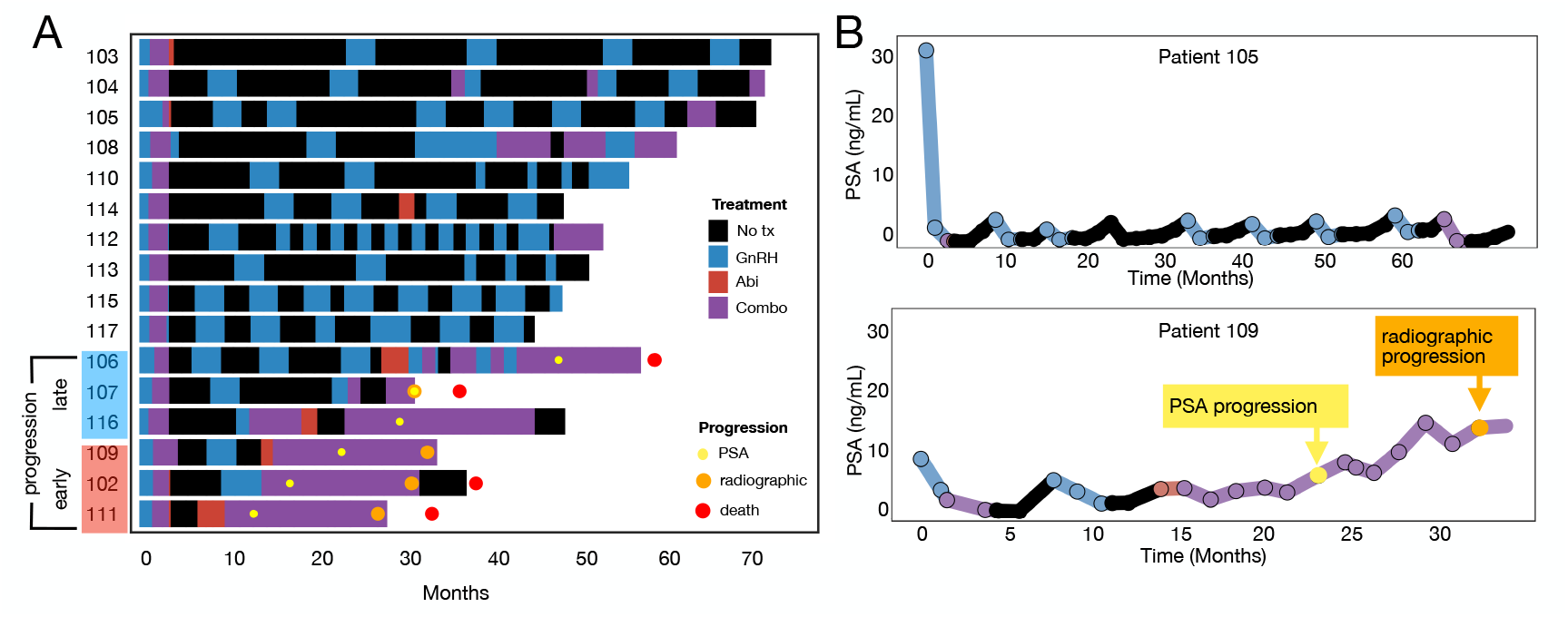
Updated trial results. A) A swimmer’s plot shows the timeline of individual patients. The early progressors (<24m) are highlighted red, whilst the late progressors are highlighted blue (>24m). The yellow, orange, and red dots indicate the time of PSA progression, radiographic progression, and death. B) PSA and testosterone dynamics are shown for a nonprogressor (patient 105) and an early progressor (patient 109).

Higher Gleason scores are generally predictive of early progression and shorter survival in metastatic prostate cancer [2], [3]. However, we have shown previously that whilst this was true for mCRPC patients in response to continuous treatment, there was no clear correlation for patients receiving adaptive therapy in which treatment is cycled on and off so that proliferation of treatment-sensitive cells is used to suppress proliferation of treatment-resistant cells [4]. A higher PSA nadir is consistently found to be predictive for progression [5], [6], [7], [8], [9], [10]. However, whilst it may be more intuitive to correlate a faster response to treatment with a better prognosis assuming that a higher proportion of sensitive cells would lead to a faster death rate [11], a longer time to PSA nadir is most often predictive of a better outcome [5], [6], [8], [10]. In advanced disease, higher testosterone levels before ADT and lower testosterone levels during ADT correlated with increased time to progression and overall survival [12].

Here we examine these and other biomarkers in the context of an evolution-informed therapy. This therapeutic strategy changes focus from killing the maximum number of treatment sensitive cells to controlling proliferation of the resistant population, which ultimately determines the clinical outcome. Thus, application of therapy is explicitly guided by evolutionary principles so that the subsequent fluctuations in the size of the treatment sensitive cancer cells result in Darwinian interactions that suppress proliferation of the resistant cells.

To identify predictive features of progression, we compare clinical features, testosterone metrics, and PSA metrics over the trial cohort. Specifically, we focus on the first cycle of adaptive therapy to identify features that may predict progression early on during the treatment course. We also fit several mathematical models to the dynamic patient-specific data to gain more mechanistic insights as to what might be driving early progression.

## Results

### Clinical trial update

While ten patients have still not progressed over the course multiple adaptive cycles, 6 patients have developed PSA progression, and 4 of these have developed radiographic progression, which is the criteria to go off trial. Four patients have died, but only the death of patient 107 at month 32.5 was attributed to progressive prostate cancer. The OS rate at month 55 is 75% in this trial. At month 48 the OS rate was 81%, which is higher than the apalutamide plus LHRH analog arm in the Titan trial (65.1% at month 48) [15] and the enzalutamide plus LHRH analog arm in the Arches trial (71% at month 48) [16]. The treatment timelines for all 16 patients are given in **Fig. 1A**. Patients spent a significant fraction of time untreated (47±20%) over the course of adaptive therapy as per the protocol schema. The earliest progressor, patient 111, had the least amount of time off treatment (11%), whilst the patient on the trial longest, patient 103, had the most amount of time off treatment (75%).

### Significant trends are found in clinical features, but there is overlap between distinct outcome groups

To better characterize features that predict progression, we categorized the cohort by time to progression (TTP), with 3 early progressors (TTP<=24 months), 3 late progressors (TTP>24 months), and 10 non-progressors (**Fig. 1A)**. Several clinical factors were analyzed to determine risk of progression. We found risk status [17], number of bone metastases, and total number of metastases were significant to distinguish PSA progressors from non-progressors with less significance in distinguishing early, late and non-progressors (**Fig. A1**). In contrast, Gleason Scores, the number of lymph node metastases, and de novo metastases were not found to be significant.

However, whilst these metrics identify significant trends, there is still significant overlap between categories so that identifying progressors with these clinical criteria is not definitive. A useful comparison that we will revisit throughout this work is between patient 105 and 109 (**Fig. 1B)**. Clinically, these patients are quite similar. They both have Gleason scores of 9, 4 identified bone metastases, and are categorized as high risk. However, whilst patient 105 has continued to cycle without progression for over 5 years and has spent 63% of their time off treatment, patient 109 had PSA progression at 20.5 months, radiographic progression at 28.3 months, and spent only 18% of their time off treatment.

### First cycle times and drug choice were not significant

Here,we analyze features and dynamics over the cohort during the first cycle of treatment, i.e. the treatment induction phase followed by the 1st regrowth phase. There were no significant trends between early,late, and non-progressors in the first cycle dynamics in general like total cycle time, the duration of the induction phase or regrowth phase,or the ratio between the two (**Fig. A2A**). Yet, some non-progressors had rather long first cycles due to very slow regrowth rates. Furthermore, the specific ARSI drug did not affect progression (**Fig. A2B**).

### Testosterone and PSA dynamics can help stratify progressors

We show that, during the treatment period, testosterone decreases until reaching some lower bound or detectability limit, where it remains generally steady whilst treatment is applied (**Fig. 2A**). During the treatment break, the testosterone rises again, sometimes with a lag, until leveling off at the baseline value. Neither baseline testosterone nor nadir testosterone were significant (**Fig. 2B**), but progressors spent less of their cycle time in physiologically normal testosterone (T>100ng/mL) (p=0.045), and progressors also had a greater percent change between the initial and final testosterone (p=0.037). All early progressors had a testosterone value at the end of the cycle that was at least 34% less than the baseline. Both features can be seen in the comparison of Patient 105 and 109 (**Fig. 2C**). Patient 105’s testosterone drops during treatment, rises again after treatment, and levels off near the baseline. Patient 109 drops quickly, lags for some time after treatment stops, and then only returns to 66% of the initial testosterone value. Categorizing between all PSA progressors and non-progressors, both metrics are significant, whereas categorizing between radiographic progressors and non-progressors only the percentage of time spent in normal testosterone conditions is significant (**Fig. A3**).

**Figure 2.**
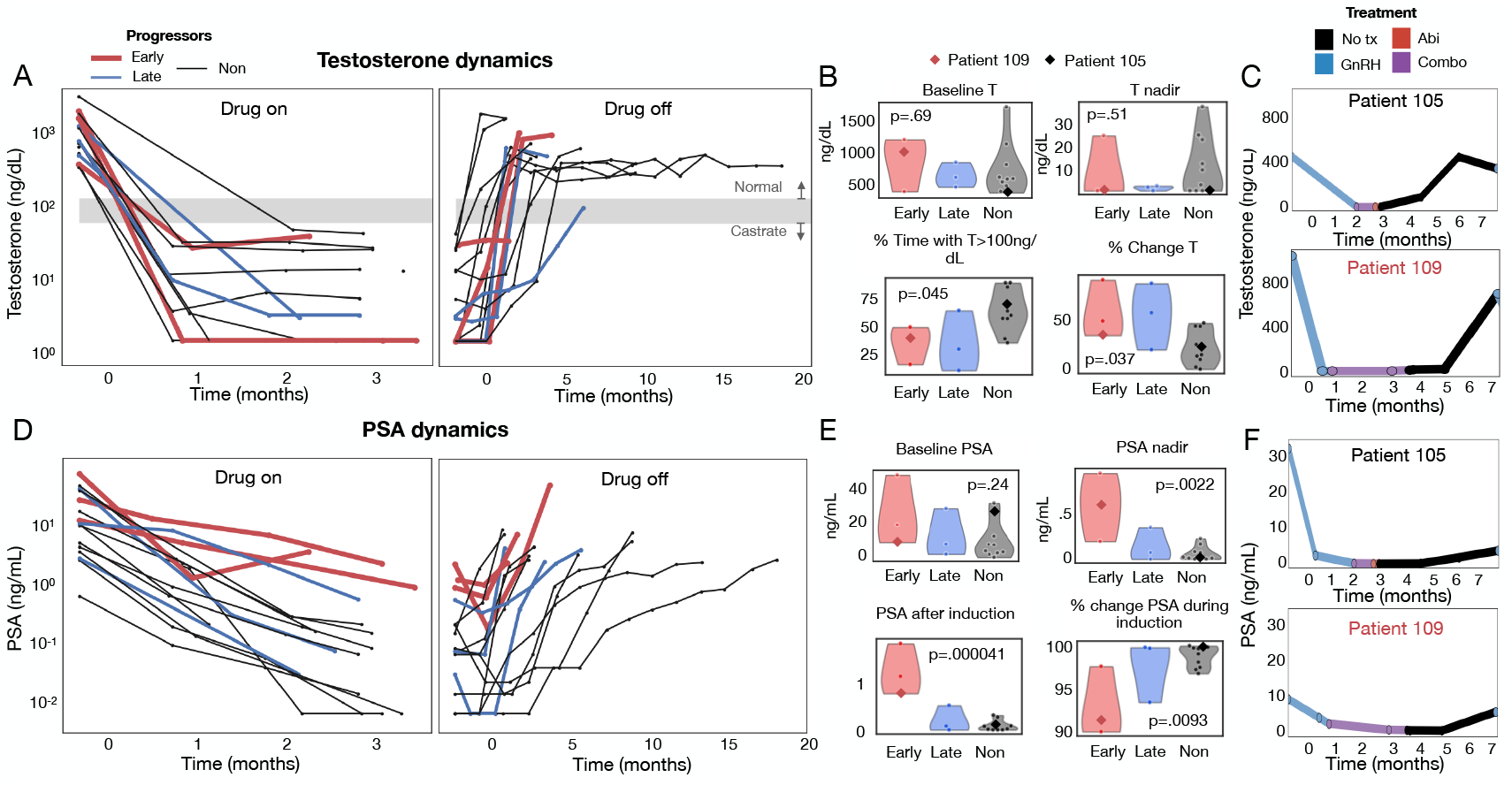
Comparing features in the dynamics of testosterone and PSA between early progressors (<24 months), late progressors (>24 months) and nonprogressors over the 1^st^ adaptive therapy cycle. A) The treatment on period (left) and treatment off period for testosterone (right). B) Baseline (initial) and the nadir testosterone values were not significant, but progressors spent less of the cycle time in normal testosterone conditions (T>100ng/mL) and had a greater % change in testosterone from baseline to the end of cycle 1. C) Comparison of testosterone dynamics between patient 105 (nonprogressor) and 109 (progressor). D) The treatment on period (left) and treatment off period (right) for PSA. E) Baseline PSA (PSA_0_) was not significant, but PSA nadir, PSA at the end of the induction phase, and the percent change in PSA during treatment were significant to discriminate progressors. Progressors tended to have a higher PSA nadir and PSA post induction and have a lower percent change in PSA during treatment. F) Comparison of PSA dynamics between patient 105 (nonprogressor) and 109 (progressor). P-values are determined by a one-way ANOVA test.

PSA features and dynamics were also analyzed over the 1^st^ cycle (**Fig. 2D**). It appears from these plots that the PSA for progressors stays higher during the treatment period and regrows quickly during the treatment holiday. This is partly confirmed with the analysis (**Fig. 2E**), as the PSA nadir and PSA post induction are both high for progressors (p=0.0022 and p=.000041, respectively) and the percent change in PSA during treatment is less for progressors (p=.0093). On the other hand, the baseline PSA, the rate of PSA change during treatment, and the rate of PSA change during the treatment break are not significant (**Fig. A4**). Whilst the progressors do not have a significantly faster PSA regrowth rate, mainly because a large proportion of non-progressing patients have long treatment off periods (**Fig. A4**). The PSA dynamics for Patients 105 and 109 reflect these significant features regardless of the large difference in initial PSA. Patient 105’s PSA goes lower quicker and rises slightly slower than Patient 109’s PSA (**Fig. 2F**). The PSA nadir, the PSA post induction, and the percentage change in PSA during treatment were significant whether we categorized between PSA progressors and non-progressors or radiographic progressors and non-radiographic progressors (**Fig. A4**).

### PSA/testosterone ratio can help stratify progressors

In the last section, several PSA and testosterone metrics were shown to be significant to discern early, late, and no progression. Here we couple PSA and testosterone, by generating metrics involving the ratio of PSA to testosterone (PSA/T), which is observed to change over the 1^st^ treatment cycle (**Fig. 3**). This ratio is somewhat higher for progressors (**Fig. 3A**). However, to determine useful metrics that predict progression, we split the first cycle into a treatment on period (**Fig. 3B left**) and a treatment break period (**Fig. 3B right**). The baseline PSA/T was not significant, however, progressors had a higher final PSA/T, average PSA/T, and maximum PSA/T ratio (**Fig. 3C**). The percent change in the ratio while treatment was on was higher for progressors and the rate of change of the ratio during the treatment break was more negative tor progressors. The most significant metrics can be better illustrated by comparing Patient 105 and Patient 109 (**Fig. 3D**). Their baseline PSA/T ratios are similar and their final PSA/T ratios are similar, but for Patient 105 PSA/T stays low and steady during the entirety of the 1^st^ cycle while for Patient 109 it rises to some maximum level while treatment is on and decreases again, leveling out during the treatment break. In this comparison, the maximum and average PSA/T are higher for Patient 109 relative to 105, whereas the other metrics are less obviously different. Not all patients have the same PSA/T dynamics, but the average and the maximum values over the 1^st^ cycle consistently distinguish progressors. If we instead categorize with PSA progression vs not, the average, maximum, and final PSA/T are the only significant metrics, and if we categorize with radiographic progression vs not, all but the final PSA/T are significant (**Fig. A5**).

**Figure 3.**
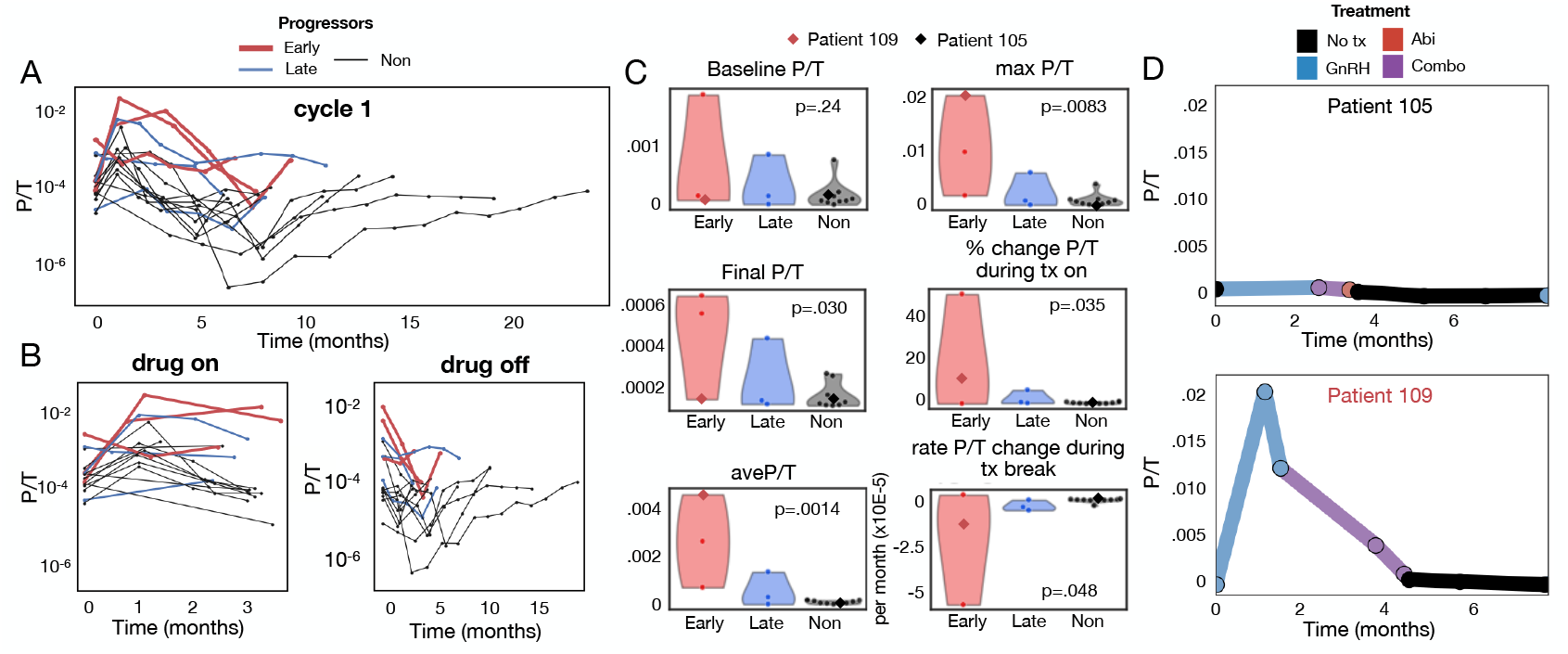
Comparing the ratio of PSA/T over the cohort. A) The ratio PSA/T for progressors appears larger over the course of the first cycle. B) PSA/T during treatment (left) and during the treatment break (right). C) Whilst the baseline PSA/T is not significant to discriminate progressors, the final PSA/T, the average PSA/T over the course of the first cycle, the maximum PSA/T, the % change in PSA/T during treatment on, and the rate of PSA/T change during the treatment break are all significant. D) The PSA/T ratio remains low over the 1^st^ cycle for nonprogressing patient 105 whilst increasing and then decreasing over the cycle for patient 109.

### A simple mathematical model shows PSA decay is slower for progressors

To better capture the growth and response trends of testosterone and PSA, we fit some simple mathematical models to the patient testosterone and PSA dynamics (**Fig. 4**). The shape of the testosterone curve is characterized as decreasing to a minimum value while on treatment and increasing to a maximum value, sometimes with a time lag, while recovering off treatment. We therefore fit the treatment-on testosterone to an exponential decay with a lower bound, and the treatment-off testosterone to a saturation function with a time delay (see **Methods** for equations). The fitted model parameters for each progression group are shown in **Fig. A6**, although no significant trends in the decay rate or the time delay parameters across the groups are observed (**Fig. 4A**). An exponential decay and exponential growth model were fit to the PSA data during the treatment on period and the treatment off period, respectively (see **Methods** and **Fig. A6**). The PSA decay rate while on treatment was just on the edge of significance (p=0.052), while the growth rate was not (**Fig. 4B**). The slower PSA response of Patient 109 relative to Patient 105 is apparent (**Fig. 4A**).

**Figure 4.**
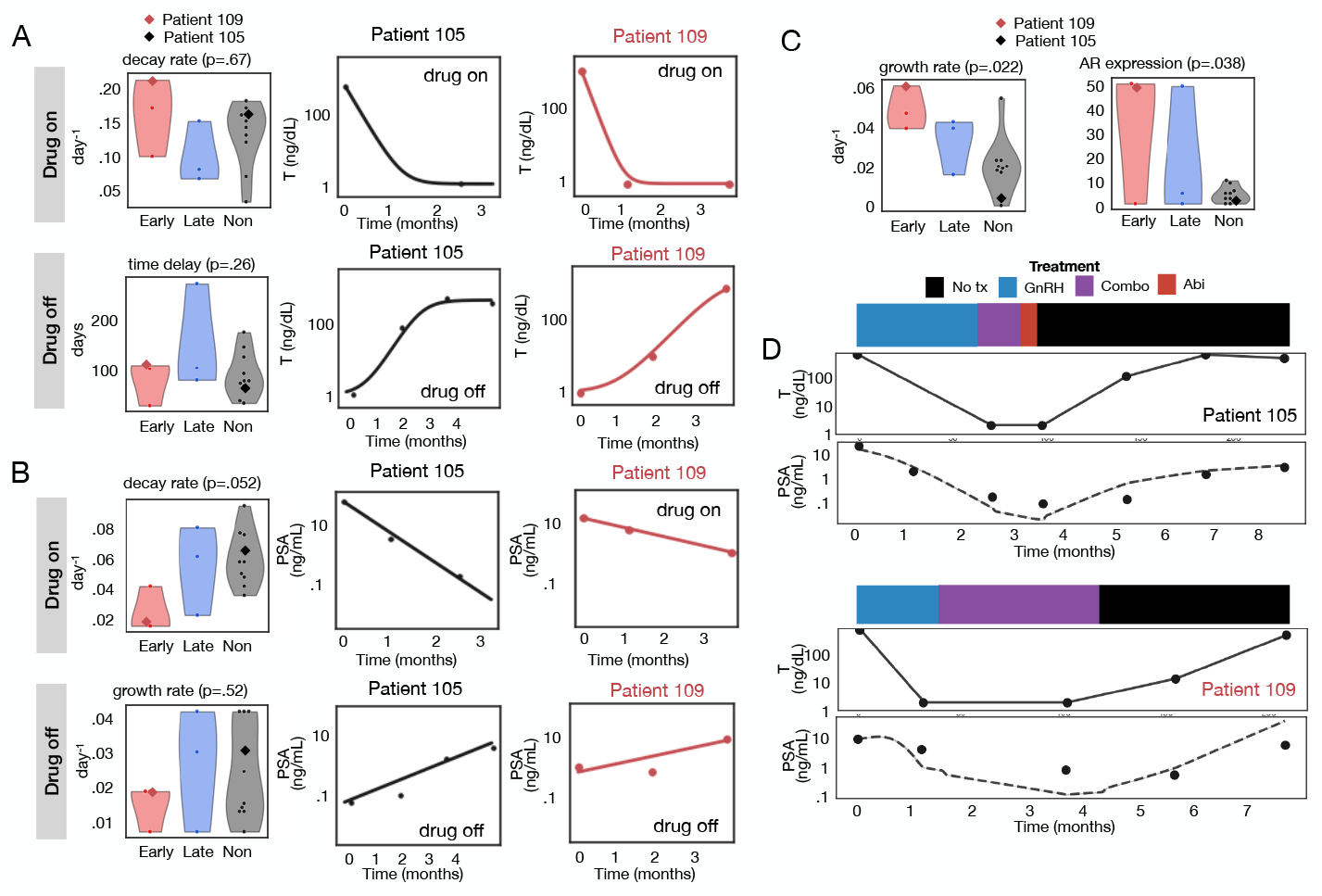
Mathematical model fits to data. A) Testosterone model fits did not reveal significant parameters. B) PSA model fits showed that the decay rate was significant, and the growth rate was not. C) For the mechanistic model, both the growth rate and the AR expression parameters were significantly higher for earlier progressors. D) The mechanistic model fits for patients 105 and 109.

### A mechanistic mathematical model suggests progressors have more resistance

We construct a simple mechanistic model that assumes the cancer population *C* dynamics consists of growth and death terms as such:

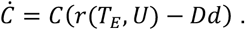

In the above equation, we assume the change in cancer over time (lefthand side) is equal to (righthand side) the cancer population multiplied by the increase due to the rate of growth *r* minus the decrease due to the rate of death *d* when an androgen deprivation drug *D* is on. The growth rate is a saturating function that depends on the effective testosterone (*T_E_=T/T_0_,* the testosterone relative to the baseline value) and the cancer’s ability to utilize the testosterone *U=AR(1-D_ARB_)*, which is increased with AR overexpression but decreased when an androgen receptor blocker *D_ARB_*is on. We fit the model to each patient’s data using their testosterone as input and assume the amount of PSA produced is proportional to the proliferation term (further details in **Methods**). Most parameters in the model are fixed whilst allowing just the growth rate *r* and androgen receptor *AR* expression to vary for each patient (see **Table A1**), we find progressors have significantly faster growth rates (p=0.022) and a significant trend (p=0.038) toward higher *AR* expression (**Fig. 4C**). The coupling of PSA with testosterone in this way can be seen by comparing Patients 105 and 109 in **Fig. 4D**. For Patient 105, the PSA dynamics almost perfectly follows the dynamics of the testosterone in lockstep, but for Patient 109, even though the testosterone plummets to its nadir quickly, the PSA lags with a slower decline, which leads to continued proliferation despite the extended reduced testosterone during treatment.

At the extreme regimes of testosterone and *AR* expression, we can simplify the PSA expression to gain intuition (see **Appendix** for derivation):

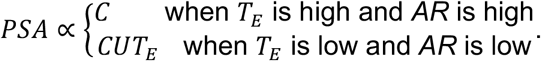

This expression states that when testosterone is high at the start and end of the adaptive therapy cycle and *AR* is high, the PSA produced is simply proportional to the number of cancer cells. However, the other extreme is when the testosterone *T_E_*goes low while *AR* is low. In this regime, there is also dependency on PSA by the amount of testosterone (reducing proliferation and therefore PSA) and a low utilization *U* via a low *AR* value. This represents a population of cells that are dependent on testosterone and respond by reducing proliferation. The other cases lie in between. A high *T_E_*and low AR leads to slightly reduced proliferation at saturation, and a high AR counteracts the low *T_E_*to allow continued proliferation and therefore production of PSA even when treatment is on.

### Cluster analysis reveals thresholds of significant features to predict progression

Several of the most discriminant features can be used together to predict progression through multivariate analysis. Looking at pairwise correlations between the PSA post induction, the average PSA/T, and the percentage change in PSA during the induction period, we find that generally 3 or 4 of the earliest progressors separate from the rest of the cohort (**Fig. 5A**). **Figure 5B** visualizes the clustering of the patients in the cohort due to these metrics using principal components, which shows that the 3 earliest progressors are clustered separately. The means of these clusters can then be used to define thresholds for predicting progression. Using the threshold for PSA post induction >.5ng/mL identifies the 4 earliest progressors. The threshold for average PSA/T >.0005 also identifies the 4 earliest progressors. The threshold for change in PSA during the induction phase <95% identifies 3 of the 4 earliest progressors.

**Figure 5.**
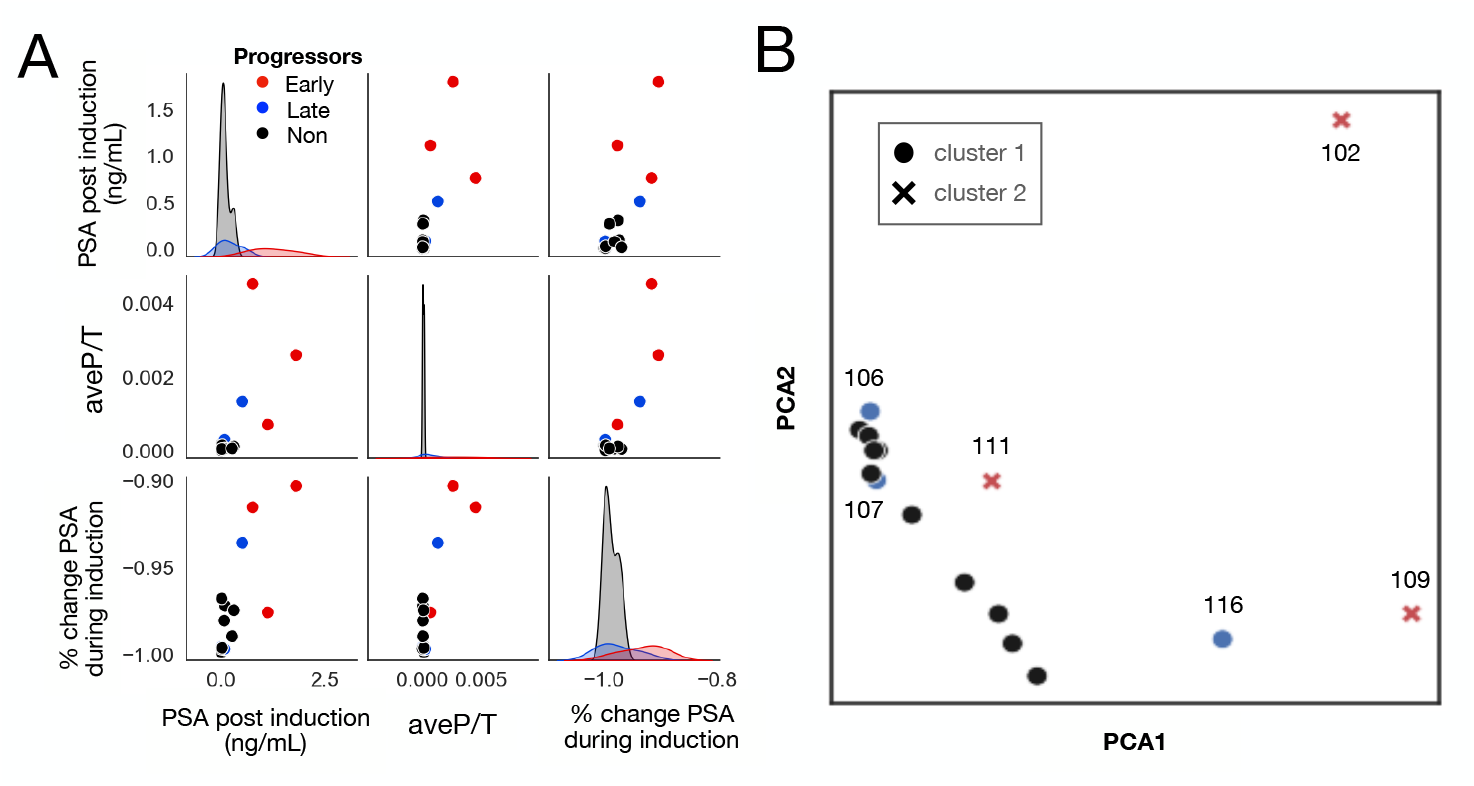
Multivariate analysis of significant predictive features. A) Distributions of and pairwise correlation between significant features. K-means cluster analysis of the same 3 features cluster the 3 earliest progressors together.

## Discussion

The ongoing trial has demonstrated that patients are, on average, receiving less than half the drug compared to continuous standard of care treatment (i.e., receiving no cancer therapy 54% of their time on trial) and the median progression free survival has not been reached at 55 months. By comparing progressors and non-progressors, we investigated potential biomarkers that predict early progression. Whilst some clinical features at baseline are correlative, they are not definitive in predicting progression, so the PSA and testosterone dynamics during the 1^st^ cycle of adaptive therapy were analyzed to gain further insights into predicting resistance.

We observed that early progressors, during the first treatment cycle (i.e., induction period plus first regrowth phase), spent less time with normal testosterone levels and testosterone often failed to return to baseline levels. Progressors had higher PSA values and lower relative change in PSA levels at the end of the induction period as well as higher PSA nadir levels, which is consistent with previous studies [5], [6], [7], [8], [9], [10]. Early progressors had higher average and maximum PSA/T ratios over the first cycle, which is in agreement with studies showing a higher PSA/testosterone ratio to be a predictor of progression during intermittent therapy [13] and a diagnostic predictor of prostate cancer in hypogonadal men [14]. Early progressors were also associated with a slower decay rate of PSA when fit to an exponential decay model and a higher AR expression when fit to a mechanistic mathematical model coupling PSA to testosterone. Collectively, these changes indicate progressors have a slower response to treatment despite testosterone dynamics that show the drug is working as intended.

Coupling the testosterone to the PSA through a PSA/testosterone ratio permits an intuitively clear understanding of the evolutionary dynamics of progression. When testosterone availability is low either in poorly vascularized regions or during ADT, prostate cancer cells can increase their fitness by: 1) increasing expression of androgen receptors or components of the AR pathway to amplify the signal despite low ligand concentrations, 2) producing androgen within the cell to generate an autocrine loop, or 3) activating the androgen receptor signaling pathway independently of ligand binding. In any case, the tumor cells will be able to better proliferate and therefore produce PSA even whilst testosterone is low. Thus, as the resistant populations increase, the serum PSA/testosterone ratio will similarly increase.

Ultimately, earlier identification of patients most at risk for progression is valuable only if it can be addressed therapeutically. Docetaxel chemotherapy has been shown to be beneficial for metastatic castrate-sensitive prostate cancer [18], and in androgen independent disease as the cytotoxic mechanisms involve proliferation pathways [19]. Other currently available agents include both cytotoxic drugs and PSMA-lutetium radioligand therapy.

An alternative approach seeks to alter the tumor composition by increasing the fitness of the ADT responsive cells and decreasing fitness of the resistant cells. This may be achieved by a transient but rapid increases in testosterone. This promotes proliferation of ADT sensitive cells and can induce apoptosis in cells with increased AR expression [20], [21], [22]. The feasibility and safety of injecting testosterone in patients with mCRPC has been demonstrated in bipolar androgen therapy (BAT), which relies on cycling between high and low levels of testosterone [23], [24]. BAT therapy is applied typically with the goal of burden reduction but our models suggest it may also be used as a form of directed evolution to restore the castrate sensitive state (mCSPC) in mCRPC tumors.

This small pilot study has its limitations. There are only 16 patients in the cohort and no explicit randomized comparison arm. However, despite these constraints significant trends were identified to predict progressors during the 1^st^ cycle of therapy using both clinical characteristics and PSA and testosterone data. Coupling the PSA and testosterone for the PSA/T ratio required more frequent sampling of testosterone than is standard, but this additional metric was essential to more precisely cluster the earliest progressors. Additionally, coupling the PSA data to the testosterone data using a mechanistic mathematical model added dynamical constraints that allowed us to map the resistance mechanisms (increased growth and AR expression) to the cancer burden (PSA) and its resources (testosterone) and gain further insight.

## Methods

### Metrics analysis

Some values for PSA and testosterone are missing over the course of treatment. If these values don’t occur at the initial time point, we ignore them, but many metrics rely on normalization by the baseline value or fall in the denominator of a ratio for which we need to fill in the values. In this case: 1) if there is a value at a time point before treatment starts, we assume that the initial value is the same, 2) if there is more than one value at other time points before treatment starts, we assume the initial value is the average of those values, and 3) if there are no pre-treatment values, which only occurs with testosterone, we assume that the initial value is equal to the maximum value during the 1^st^ cycle. This assumption is based on testosterone returning to its baseline off treatment.

For the statistical analyses, we used the one-way ANOVA test in Python to find p-values to distinguish 3 groups: early, late, and nonprogressors, or 2 groups PSA progressors vs nonprogressors or radiographic progressors vs non-radiographic progressors. For the cluster analysis we used the k-means clustering algorithm from the scikit-learn package in Python.

### Mathematical models

#### Models for testosterone

We fit the data separately to testosterone *T* over time *t* during the treatment phase and the regrowth phase when treatment is off. The former was fit to an exponential decay that levels off at a minimum value:

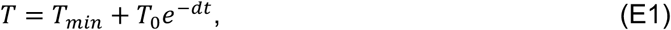

where *T_min_*is the lower bound of testosterone during treatment, *T_0_*is the baseline testosterone, which are derived from the data, while *d* the decay rate is fit. Testosterone during the growth phase was fit to a time-delayed growth saturation model:

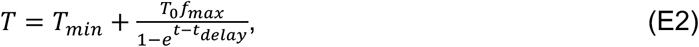

where *f_max_*is fraction of the baseline testosterone that is achieved during the regrowth phase, and *t_delay_*which are both fit to each patient.

#### Exponential models for PSA

The PSA was fit separately to the data during the treatment phase and the regrowth phase when treatment is off. The PSA during the treatment on phase was fit to an exponential decay function:

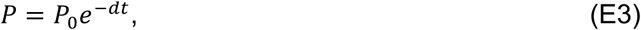

where *P_0_*is the initial PSA, and *d* is the death rate. PSA during the treatment off phase was fit to an exponential growth function:

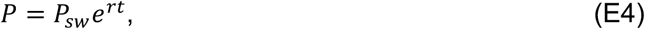

where *P_sw_*is the PSA at the treatment switch off time point, and r is the growth rate. For these equations, *P_0_*and *P_sw_*are derived from the data, whilst *d* and *r* are fit.

#### Mechanism-based ODE model

We construct a simple ordinary differential equation (ODE) model to account for growth and death dynamics of prostate cancer cells *C*:

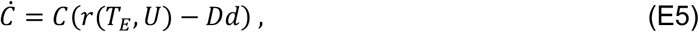

where the rate of change in cancer burden *C* is equal to the burden multiplied by the increase due to rate of growth *r* minus the decrease due to the rate of death *d* when an androgen deprivation drug *D* is on (D=1 when LHRH and abiraterone are on, D=.8 when LHRH is on, and D=.5 when abiraterone is on). The growth rate *r* depends on the effective testosterone (*T_E_=T/T_0_,* the testosterone relative to the baseline value) and the cancer’s ability to utilize the testosterone *U=AR(1-D_ARB_)*, which is increased with AR overexpression *AR* but decreased when an androgen receptor blocker *D_ARB_*is on. The growth rate function is as such:

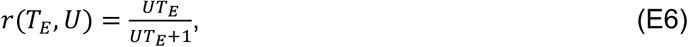

which is low with low testosterone and saturates at high values. We assumed that the amount of PSA produced is proportional to the proliferation rate of the cells, i.e. the first term in E1 and the production rate of PSA *r_PSA_*from each cell. However, using the assumption that *C* is equal to PSA at time 0, the *r_PSA_*and r cancel out:

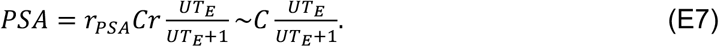

To solve this model, we linearly interpolate the patient-specific testosterone values to be used as input during fitting. We minimize the error between the model fits and data points to get the optimal *r* and *R*. All other parameters are global and are listed in **Table A1**.

## Data Availability

All data produced in the present study are available upon reasonable request to the authors.

## Acknowledgments of Research Support

The authors gratefully acknowledge funding from the National Cancer Institute via the Cancer Systems Biology Consortium (CSBC) (U54CA274507) and support from the Moffitt Center of Excellence for Evolutionary Therapy.

## Appendix

**Table A1.** Parameters used for the mechanistic mathematical model.

| Description | Name | Value |
| --- | --- | --- |
| Growth rate | $r$ | 0.001-0.07 |
| Death rate | $d$ | 0.03 |
| Response to AA | $D_{AA}$ | 0.25 |
| Resistance factor | $U$ | 1-50 |

## Derivation of PSA approximation

We start with equation E7

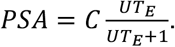

When *T* is high, *T_E_*approaches 1, so

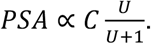

When *U* is also high via a high *AR, U/(U+1)* approaches 1, so PSA*∼C*. However, when *T_E_*is low and *U* is low, we can assume

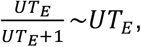

by using the Maclaurin series approximation where x/(1+x) approaches x when x is small. And in the case that *T_E_*is low and *U* is high we cannot cleanly simplify any further. To sum up,

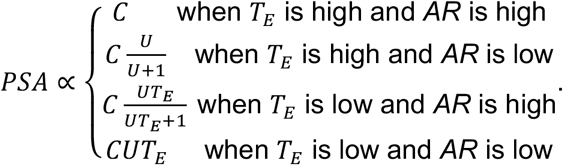

**Figure A1.**
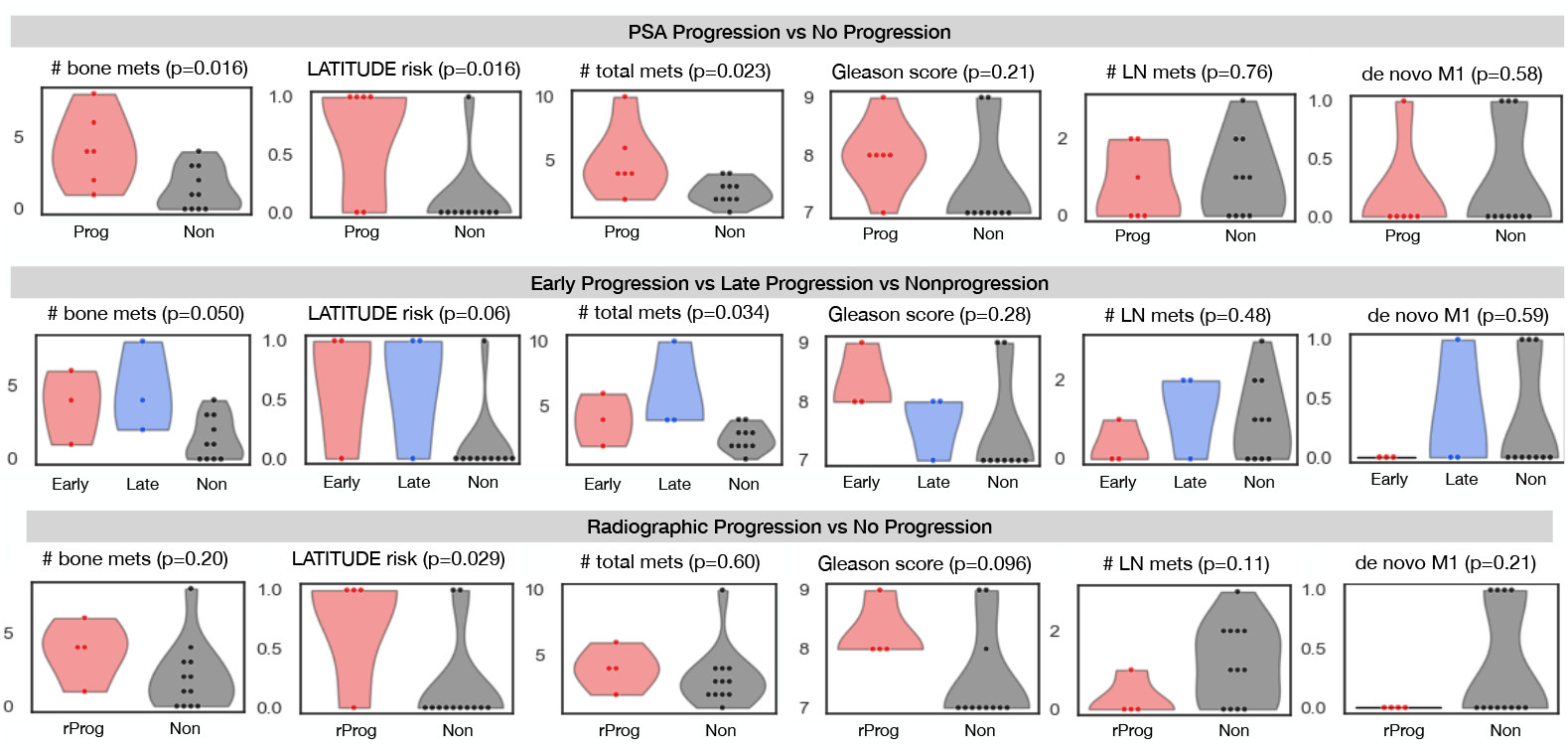
Comparing clinical features between PSA progressors and nonprogressors (top), early (<24 months), late (>24 months), and nonprogressors (middle), and radiographic progressors and nonprogressors. PSA progression vs nonprogressors could be distinguished significantly with number of bone metastases, risk status, and total # metastases. Early, late, and nonprogressors could be distinguished significantly with numer of bone metastases and total number of metastases. Radiographic progressors and nonprogressors could be distinguished significantly with risk status only. Significance was determined using a one-way ANOVA test with p-value<=0.05.

**Figure A2.**
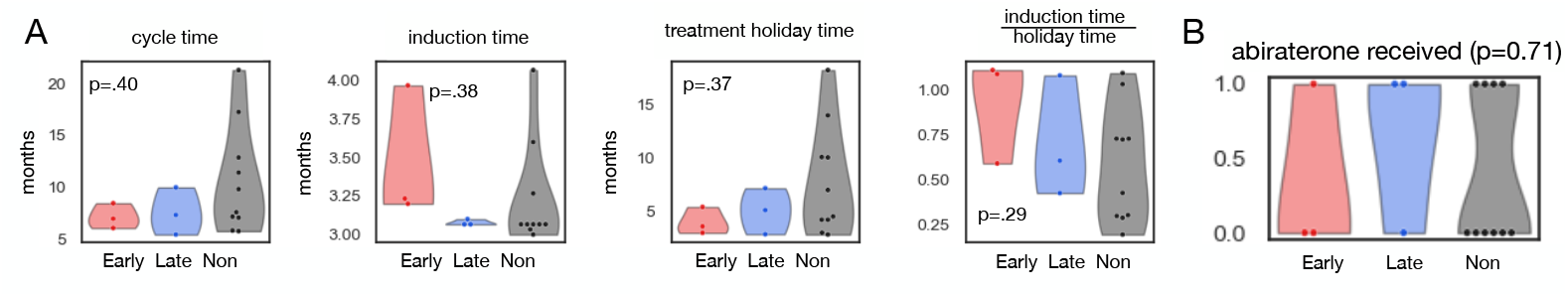
A) Cycle timing was not significant to determine progression group. B) Receiving abiraterone or some other ARSI was not significant to determine progression group.

**Figure A3.**
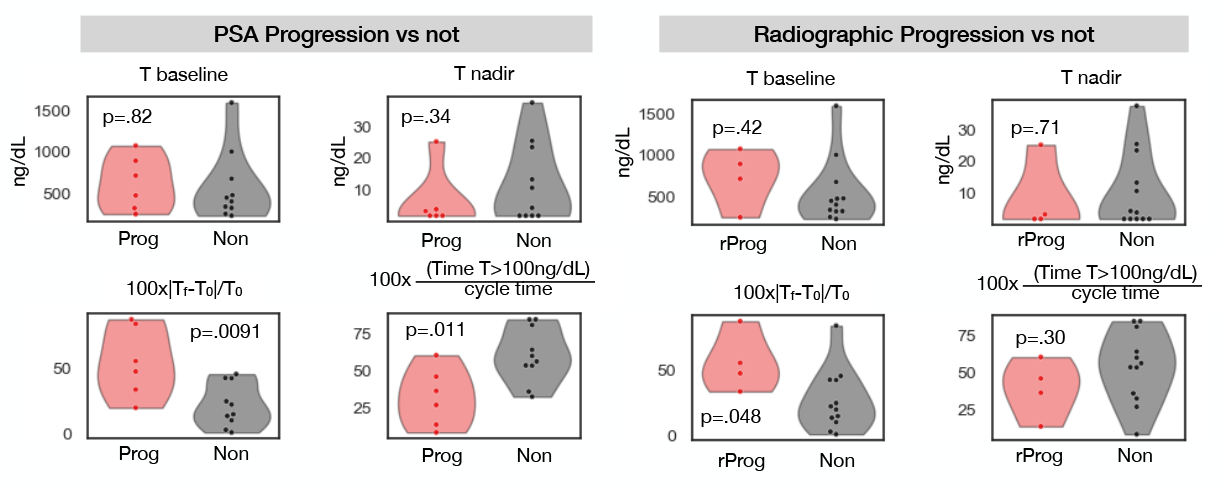
Testosterone metrics comparison. A greater percent change in testosterone was significant to discriminate PSA and radiographic progressors from nonprogressors. Less time spent in normal testosterone (T>100ng/mL) within the cycle was significant to discriminate PSA progressors from nonprogressors.

**Figure A4.**
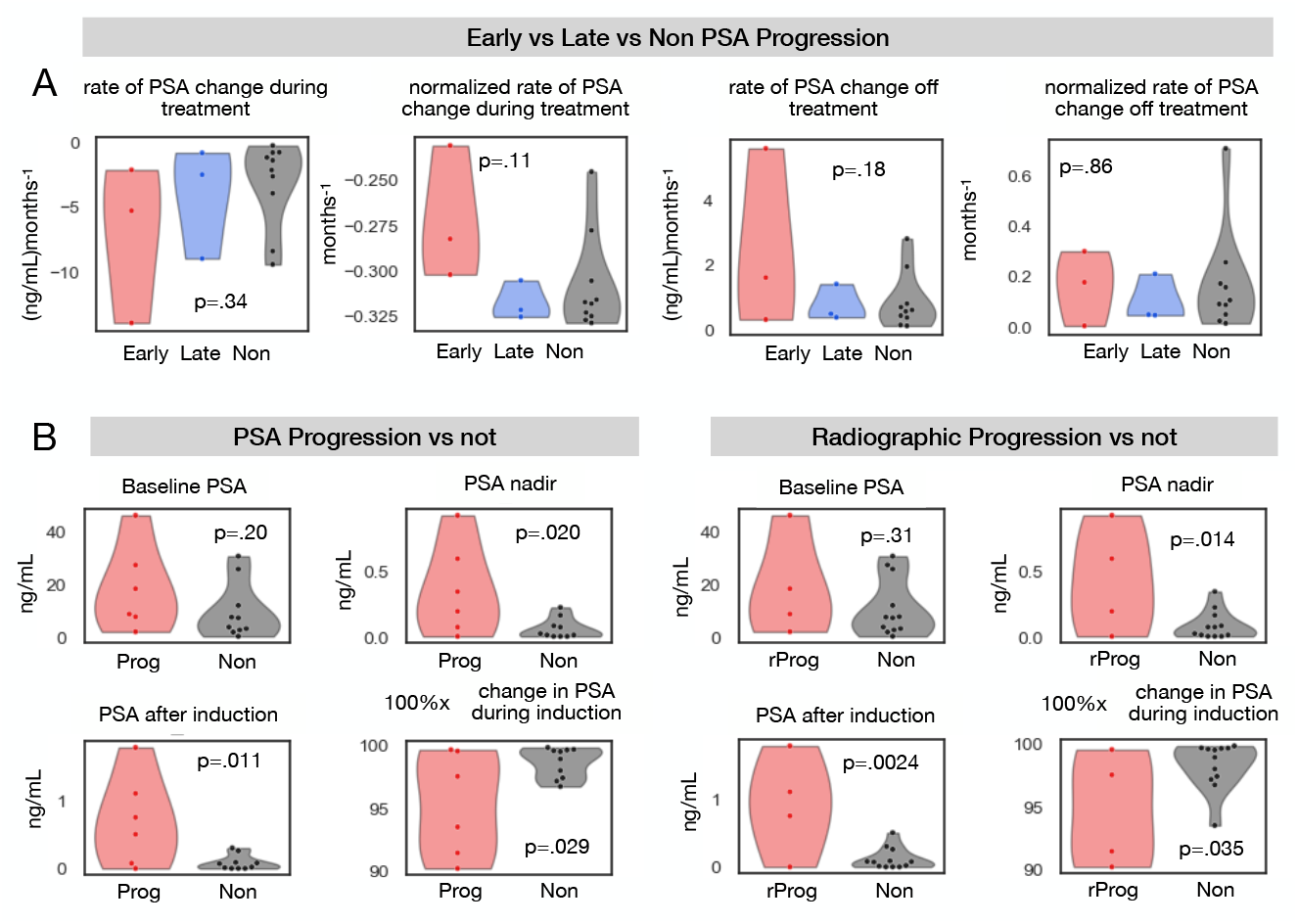
Metrics describing PSA: A) rates of PSA change, and B) metrics from Fig. 2E categorized instead between whether the patients had PSA progression or not (left), and whether they had radiographic progression or not (right).

**Figure A5.**
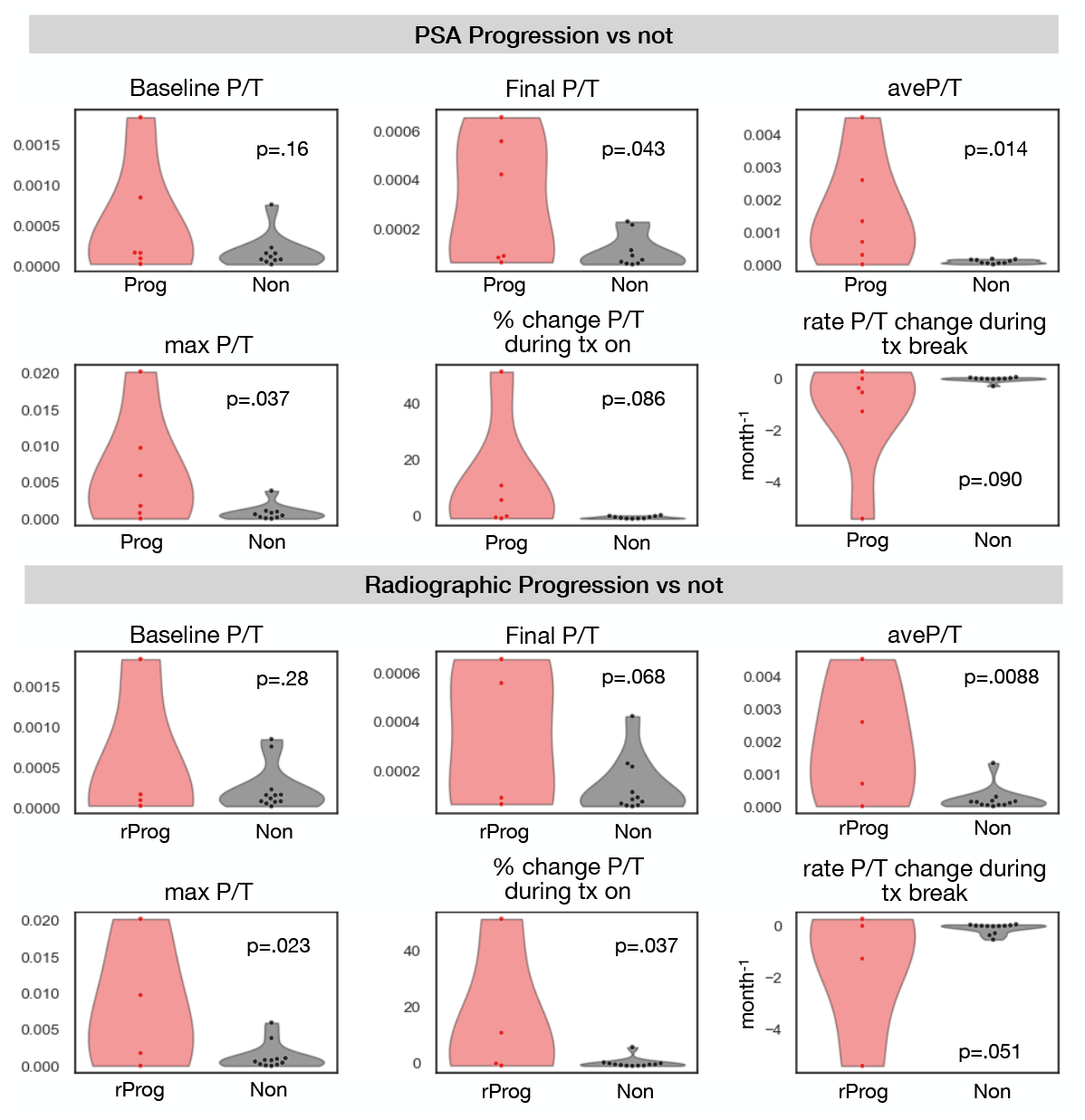
PSA/Testosterone ratio metrics comparing PSA progressors and nonprogressors (top) and radiographic progressors vs nonprogressors (bottom).

**Figure A6.**
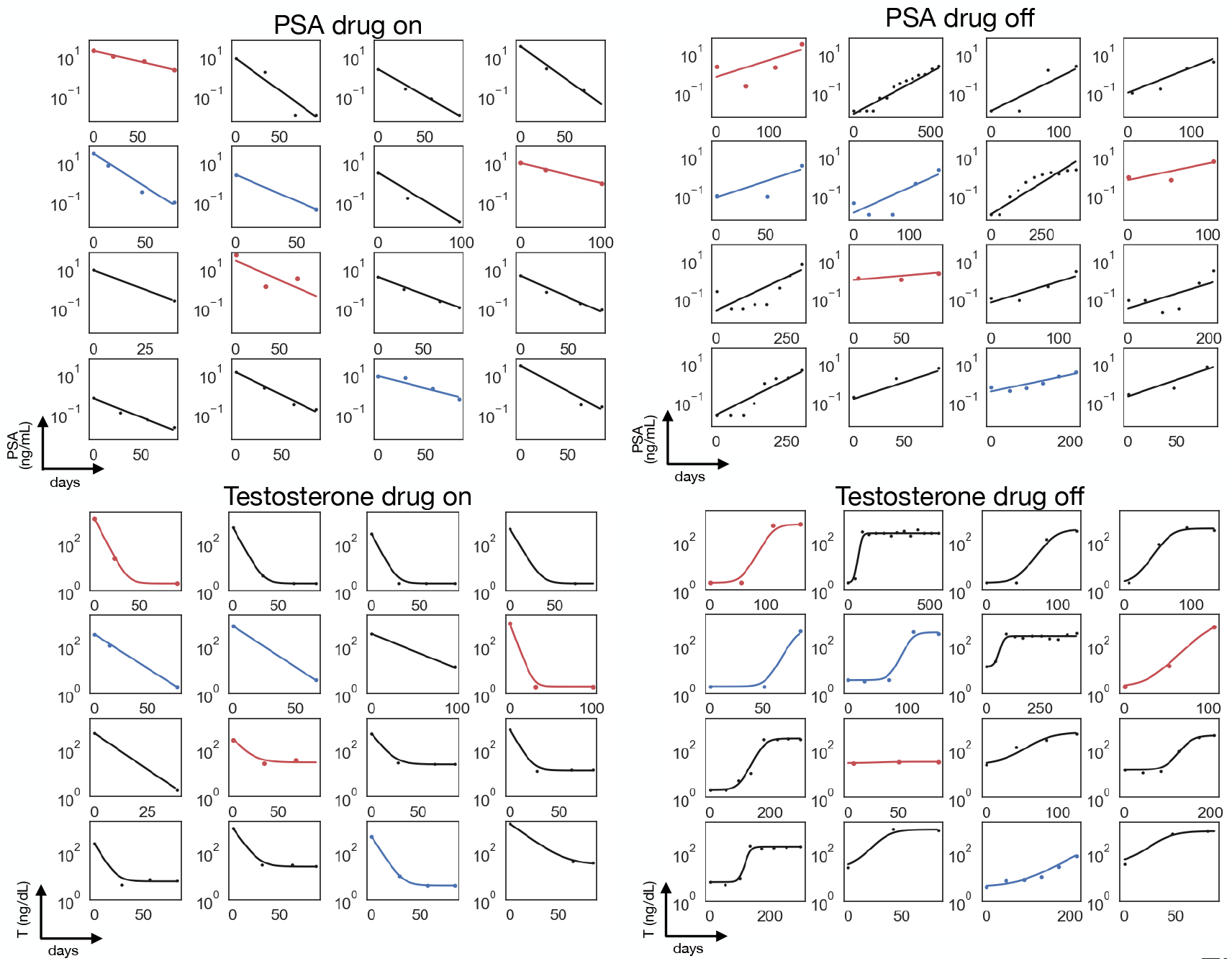
PSA and testosterone dynamics were fit to the functions described in the **Methods** section.

